# Initial treatment choices for long term remission of insomnia disorder in adults: a systematic review and network meta-analysis

**DOI:** 10.1101/2024.04.24.24306311

**Authors:** Yuki Furukawa, Masatsugu Sakata, Toshi A. Furukawa, Orestis Efthimiou, Michael Perlis

## Abstract

**Background:** Cognitive behavioral therapy for insomnia (CBT-I), pharmacotherapy and their combination are effective for insomnia. However, it remains unclear which treatment is more likely to lead to favorable long-term outcomes when used as the initial treatment. We aimed to evaluate the comparative efficacy and acceptability of CBT-I, pharmacotherapy, and their combination in the long- and short-terms among adults with insomnia disorder.

**Methods:** We searched PubMed, CENTRAL, PsycINFO and WHO ICTRP from database inception to Dec 27, 2023, to identify published and unpublished randomized controlled trials. We included trials in hypnotic-free adults with insomnia disorder comparing at least two of the following: CBT-I with at least one effective component (sleep restriction, stimulus control, cognitive restructuring, and third wave components), pharmacotherapy, or their combination. We assessed the confidence in evidence using CINeMA. The primary outcome was long-term remission (longest follow-up between 3 to 12 months). Secondary outcomes included all-cause dropout and self-reported sleep continuity measures at long-term follow-up, and the same outcomes at the end of the acute treatment phase. We performed frequentist random-effects network meta-analyses. We used odds ratio (OR) for dichotomous outcomes and mean difference for continuous outcomes, expressed in minutes and percent. This study is registered in PROSPERO (CRD42024505519).

**Findings:** We identified 13 trials, including 823 randomized participants (mean age, 47.8 years, 60% women). Results suggested that CBT-I was more beneficial than pharmacotherapy in the long-term (remission OR 1.82 [95% Confidence Interval (CI), 1.15 to 2.87; certainty of evidence: high]), while there was weaker evidence of benefit of combination against pharmacotherapy (OR 1.71 [95%CI, 0.88 to 3.30: moderate]) and no clear evidence of difference of CBT-I against combination (OR 1.07 [95%CI, 0.63 to 1.80: moderate]). CBT-I was associated with less dropouts than pharmacotherapy in the long-term. Short-term outcomes also favored CBT-I over pharmacotherapy except total sleep time. Given the average long-term remission rate in the pharmacotherapy-initiating arms of 28%, CBT-I resulted in a long-term remission rate of 41% (95% CI: 31% to 53%) and combination 40% (95% CI: 25% to 56%).

**Interpretation:** This study found that starting with CBT-I for the treatment of adults with chronic insomnia leads to better outcomes than starting with pharmacotherapy. Combination therapy may be better than pharmacotherapy alone, but unlikely to be worth the additional burden over CBT-I alone.

**Funding:** None.

## INTRODUCTION

Insomnia is common and disabling.[1] As many as 8% of the people in the United States use sleep medications in 2020[2], with this rate having doubled in the last decade.[3] Around 20% of hypnotic users are prescribed sleeping aids for longer than 180 days.[4,5] This is concerning, given that a recent network meta-analysis (NMA) found very sparse evidence supporting hypnotics in the long-term.[6] Another treatment option is cognitive-behavioral therapy for insomnia (CBT-I), a non-pharmacological intervention that is now recommended as the first-line treatment[7] and has been shown to be effective in the long-term.[8] Although many patients prefer non-pharmacological treatments over medications, non-pharmacological options are rarely provided.[9] Factors impeding the dissemination of CBT-I includes not only the lack of clinicians’ confidence in administering it but also the lack of knowledge among clinicians and patients regarding its comparative efficacy against pharmacological therapies.[10]

Another recent NMA suggested potential superiority of CBT-I over pharmacotherapies (sleeping medications) and superiority of combination therapy (CBT-I plus pharmacotherapies) over pharmacotherapies alone, for people with insomnia with or without sleeping medications at the end of the acute phase treatment.[11] However, this NMA included hypnotic-resistant insomnia and hence could not answer the clinical question of which treatment strategy to choose when starting to treat medication-naïve insomnia. Moreover, it had important methodological limitations, such as violation of transitivity assumption (e.g., hypnotic users were included for pharmacotherapy vs combination comparison, but excluded for comparisons including psychotherapy alone arm), including non-pharmacological interventions not shown effective for insomnia (e.g., sleep hygiene education and relaxation)[12], and including medications not normally used for treating insomnia (e.g., dexmedetomidine). Also, it could not provide conclusions about the long-term comparative efficacy.

In this study, we explored the long-term relative efficacy and acceptability of CBT-I, pharmacotherapy, and their combination as the initial treatment choice with the use of NMA, focusing on trials that randomized people not currently on treatment for their insomnia.

## METHODS

We followed the Preferred Reporting Items for Systematic reviews and Meta-Analyses (PRISMA) guideline extension for NMA.[13] This protocol is prospectively registered in PROSPERO (CRD42024505519) and can be found in eAppendix1.

## Data sources

### Criteria for considering trials for this review

We included all randomized controlled trials that compared CBT-I, pharmacotherapies, or their combination against each other in the treatment of hypnotic-free adults with chronic insomnia. We included trials of patients of both genders aged 18 years or older with insomnia either diagnosed according to formal diagnostic criteria (such as the Diagnostic and Statistical Manual of Mental Disorders, the International Classification of Diseases, or the International Classification of Sleep Disorders) or judged so by clinical experts (e.g. presence of significant symptoms). The criteria needed to include significant distress or daytime impairment. We tested the effect of including studies without a formal diagnosis of insomnia in a sensitivity analysis. We included patients with psychiatric or physical comorbidities. We excluded trials if patients currently using prescription or over-the-counter sleep medications were included. We excluded trials focusing on insomnia not responsive to psychotherapy or pharmacotherapy for insomnia, but included trials if patients discontinued the medications for a certain period before randomization. We regarded CBT-I as a psychotherapy involving any one of the following components shown effective in a recent component NMA [12]: sleep restriction, stimulus control, cognitive restructuring, and third wave components (mindfulness and acceptance and commitment therapy (eAppendix1). We included drugs that were proven to be effective in a recent NMA [6] (benzodiazepines, doxylamine, eszopiclone, lemborexant, seltorexant, suvorexant, trazodone, zaleplon, zolpidem, zopiclone). Where multiple arms were reported in a single trial, we included only the relevant arms.

### Search methods for identification of studies

We carried out a comprehensive literature search in PubMed, CENTRAL and PsycINFO from database inception to 27^th^ December 2023. We used a combination of index and free terms of psychological and pharmacological treatments and insomnia with filters for randomized clinical trials (eAppendix1). We also searched WHO International Clinical Trials Registry Platform. We imposed no date, language, or publication status restriction at the search stage, but we included only trials in English at the screening stage. We checked the reference lists of identified studies and review articles for additional potentially eligible records.

## Data collection and analysis

### Selection of studies

Two review authors (YF and MS) independently screened titles and abstracts of all potential studies we identified in our systematic search. We retrieved the full-text study reports, and two review authors independently screened the studies for inclusion and recorded reasons for exclusion of the ineligible studies. We resolved any disagreement through discussion. We identified publications from the same study so that each study rather than each report is the unit of analysis in the review. We assessed the inter-rater reliability of the full text screening decisions with Cohen’s κ and percentage agreement.

### Data items

Two review authors (YF and MS) extracted independently data from the included studies. Any disagreement was resolved through discussion. We assessed included trials using the revised risk of bias tool by Cochrane.[14] Any disagreement was resolved through discussion. We measured the inter-rater reliability of the overall risk of bias assessment with Cohen’s κ and percentage agreement, and that of the extracted primary outcomes with intra-class correlation.

### Primary outcome and secondary outcomes

The primary outcome of interest was treatment remission, defined as reaching a satisfactory state, measured by any validated self-reported scale (e.g. the Insomnia Severity Index 7 or lower; the Pittsburgh Sleep Quality Index 5 or lower; sleep efficiency 85% or more; sleep latency 30 minutes or shorter) at long-term follow-up (longest follow-up between 3 to 12 months). We prioritized intention-to-treat analyses whenever possible. When original publications did not report the number of remitters, we imputed remission based on continuous outcomes using a previously validated method.[12,15] Secondary outcomes included all-cause dropouts (as a proxy measure of acceptability), various self-reported sleep continuity measures, including sleep efficiency (%), total sleep time (minutes), sleep latency (minutes) and wake after sleep onset (minutes). We also examined short-term outcomes (outcomes at post-treatment of the first-step treatment phase). We used odds ratios for analyzing dichotomous outcomes[12,16]. We translated the odds ratios into the experimental event rates using the weighted mean proportion of remitters in the pharmacotherapy-initiating arms as the control event rate, aiming to improve interpretability.[17,18] We used mean difference for continuous outcomes expressed in minutes and percentages, and standardized mean differences for continuous outcomes measured in variable scales.

### Statistical analysis

We created a network diagram to visualize the available evidence. Transitivity is a fundamental assumption behind NMA.[19] Transitivity implies that we can combine the direct evidence from A vs C and B vs C studies to learn indirectly about the comparison A vs B. This, however, will be questionable if there are important differences in the distribution of the effect modifiers across treatment comparisons. To assess transitivity, we created box plots of trial and patient characteristics deemed to be possible effect modifiers (publication year, age, and baseline severity) and visually examined whether they were similarly distributed across treatment comparisons. We checked consistency using global (design-by-treatment) and local (back-calculation) tests.[20,21] Given the expected variability in the patients and treatments to be included, we conducted a random-effects NMA. We visualized NMA results using pharmacotherapy as reference and ordering treatments according to p-scores, which provide an overall ranking of treatments.[22] We assessed heterogeneity by looking at the standard deviation of random effects (τ^2^) and comparing it against empirical distributions,[23] and by creating prediction intervals.[24] We assessed possible reporting bias and small-study effects using contour-enhanced funnel plots when ten or more trials were available for a single comparison. We assessed certainty of evidence using CINeMA.[25] We performed pre-specified sensitivity analyses on the primary outcome to examine the influence of including studies with informal diagnostic criteria, comorbidities, high dropout rates, and high overall risk of bias. In addition, we conducted post hoc sensitivity analyses categorizing arms using both the initial treatment but also the second-step treatment, one with a broad definition (CBT-I, combination, and pharmacotherapy) and another with a stringent definition (e.g., acute CBT-I followed by post-acute combination therapy, acute pharmacotherapy followed by CBT-I, etc…).

We conducted analyses in *R*[26] using *netmeta*,[27] and *meta*[28] packages.

## RESULTS

We identified 560 records for the title and abstract screening, then assessed 111 full-texts. We included 9 trials and 627 participants for the primary outcome (long-term follow-up) and 13 trials with a total of 823 participants for the post-treatment assessment (eAppendix2). The inter-rater reliability of judgements for full text screening was substantial, with κ of 0.65 (95% confidence interval [CI], 0.51-0.79) and percentage agreement of 84%.

Typical participants were middle-aged with moderate insomnia symptoms (mean [standard deviation (SD)] age, 47.8 [13.5] years, based on 12 trials; 510 of 823 [62%] female, reported in 13 trials; baseline Insomnia Severity Index score, 17.4 [4.0], reported in 6 trials). 12 trials used formal operationalized criteria. 9 trials had 2 arms and 4 trials had 3 arms according to our categorization (11 CBT-I-initiating arms, 8 combination-initiating arms, 11 pharmacotherapy-initiating arms). Of the 19 arms including CBT-I components, 18 arms included stimulus control, 17 sleep restriction, 15 cognitive restructuring and 1 third wave components. Of 19 arms including pharmacotherapy, 6 arms used zolpidem, 4 temazepam, 4 zopiclone, 2 trazodone, 2 triazolam and 1 eszopiclone. 9 trials reported outcomes at long-term follow-up (median duration 24 weeks, range 12 to 48 weeks) and 13 trials those at post-treatment (median 8 weeks, range 2 to 12 weeks). Table 1 shows the characteristics of the included trials. Interrater reliability of extracted primary outcomes was almost perfect, with an intraclass correlation of 0.95 (95% CI, 0.92-0.97). The overall risk of bias for the primary outcome according to the Cochrane revised risk of bias tool was low in 2 of 9 trials (22%), some concerns in 4 (44%), and high in 3 (33%) The interrater reliability for the overall risk of bias was moderate, with a squared weighted κ of 0.41 (95% CI, 0.03-0.78) and percentage agreement of 33%.

**Table 1.** Characteristics of the included trials.

|  | Value | Trials |
| --- | --- | --- |
| Age, mean (SD), y | 47.8 (13.5) | 12 |
| Sex, No. (%) |  |  |
| Female | 510/823 (62%) | 12 |
| Male | 313/823 (38%) | 12 |
| Baseline severity (ISI score), mean (SD) | 17.4 (4.0) | 6 |
| Diagnosis |  |  |
| Formal operationalized criteria |  | 12 |
| Others |  | 1 |
| Region |  |  |
| North America |  | 8 |
| Europe |  | 3 |
| Asia |  | 2 |
| Publication year, mean (range) | 2009 (1993 - 2020) | 13 |
| No. of arms |  |  |
| Total | 30 Arms | 13 |
| CBT-I-initiating | 11 Arms | 11 |
| Combination-initiating | 8 Arms | 8 |
| Pharmacotherapy-initiating | 11 Arms | 11 |
| Delivery method |  |  |
| Individual |  | 8 |
| Group |  | 5 |
| Self-help |  | 0 |

**Table 2.**
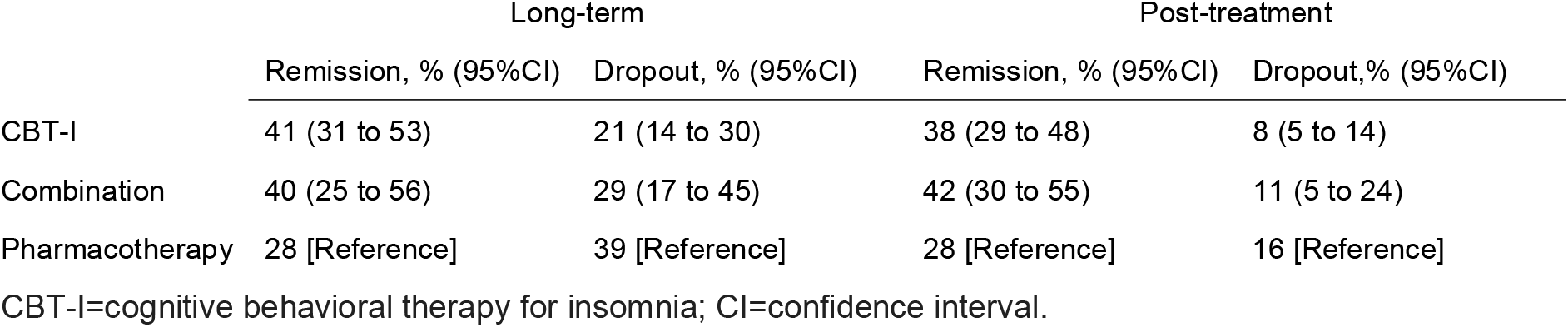
Estimated event rates for each condition.

Figure 1 shows the network for the primary outcome and Figure 2 the result of network meta-analysis. eAppendix3 shows the assessment of transitivity, which found that potential effect modifiers were evenly distributed across comparisons. The global (design-by-treatment) test showed some evidence of inconsistency (p = 0.06), but the local (back-calculation) method did not find disagreements between direct and indirect comparisons.(eAppendix3) Heterogeneity of the primary outcome was limited (T^2^ = 0.02), which was smaller than the majority of the existing meta-analyses of mental health indicators comparing non-pharmacological interventions against pharmacological interventions.[23] There was weak evidence of discrepancies between direct and indirect comparisons for CBT-I vs combination and CBT-I vs pharmacotherapy comparisons, but the indirect estimates were imprecise and the prediction intervals incorporating inconsistency did not meaningfully change the overall interpretation of results. (eAppendix3) We did not evaluate publication bias and small-study effects using funnel plots due to the limited number of trials. Figure1 shows the result of NMA for the primary outcome and eAppendix4 shows the results of the the league tables. eAppendix6 shows the result of CINeMA for the primary outcome.

**Figure 1.**
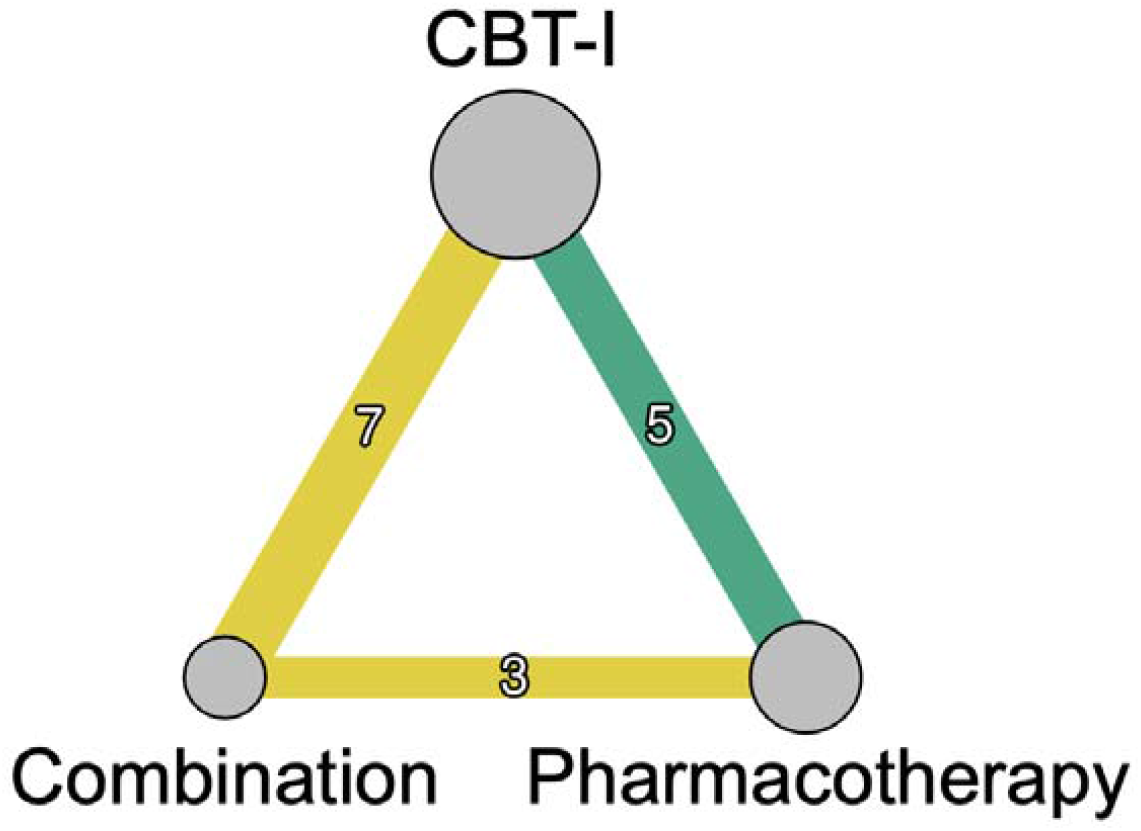
Network diagram for the primary outcome. The size of the nodes corresponds to the number of participants randomized to the treatment. The width of lines connecting treatments corresponds to the number of trials. This number is also shown on each line. CBT-I=cognitive behavioral therapy for insomnia. Colors indicate the confidence in the evidence: green=high, yellow=moderate.

**Figure 2.**
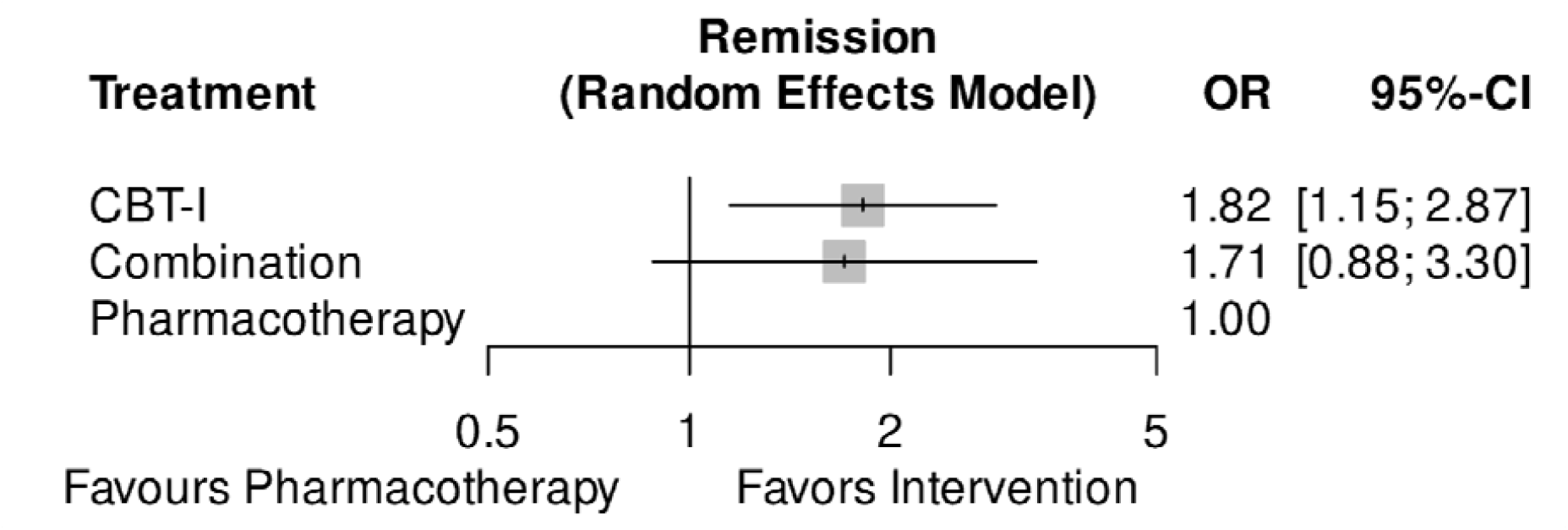
Result of network meta-analysis for remission in the long term. CBT-I=cognitive behavioral therapy for insomnia; CI=confidence interval; OR=odds ratio.

Figure 3 tabulates the results of network meta-analyses for the primary and secondary outcomes. We applied the Kilim plot,[29] coloring cells in shades of green and red, according to the strength of statistical evidence against the null. We found evidence that initiating the treatment with CBT-I (9 arms, n=292) was more effective than with pharmacotherapy in the long-term (7 arms, n=193) (odds ratio [OR] 1.82 [95%CI, 1.15 to 2.87; certainty of evidence: high]). We also found weaker evidence of superiority of combination over pharmacotherapy alone (OR 1.71 [95% CI, 0.88 to 3.30; moderate]). We did not find evidence of superiority of CBT-I over combination (5 arms, n=142) (OR 1.07 [95%CI, 0.63 to 1.80: moderate]).

**Figure 3.**
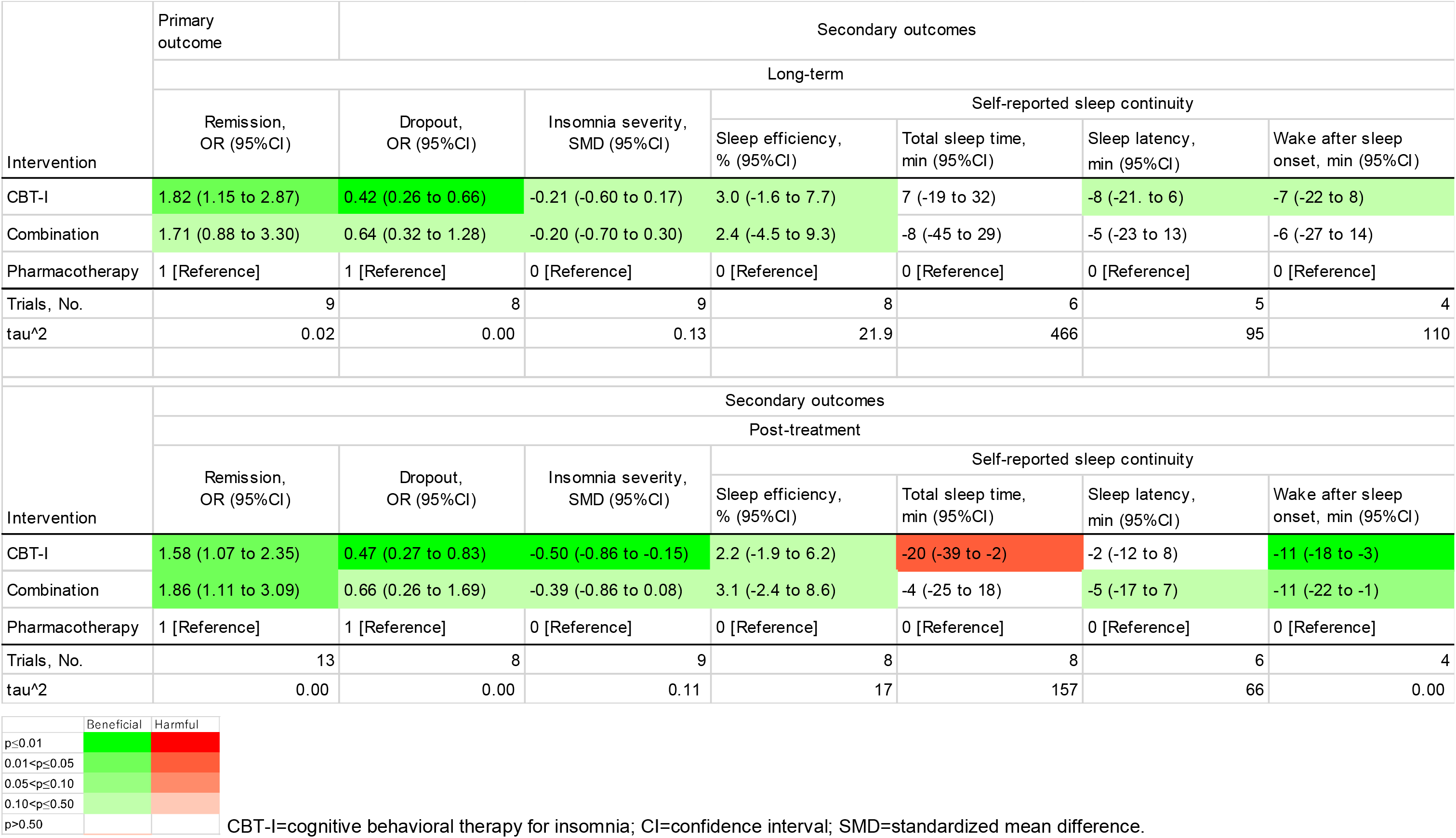
Results of network meta-analyses for primary and secondary outcomes.

CBT-I was more beneficial than pharmacotherapy in various secondary outcomes; dropout in the long-term (OR 0.42 [95%CI, 0.26 to 0.66]), remission at post-treatment (OR 1.58 [95%CI, 1.07 to 2.35]), dropout at post-treatment (OR 0.47 [95% CI, 0.27 to 0.83]), insomnia severity at post-treatment (standardized mean difference [SMD] −0.50 [95%CI, −0.86 to −0.15]), and wake after sleep onset at post-treatment (mean difference [MD] −11 minutes [95% CI, −18 to −3]). However, CBT-I led to shorter total sleep time at post-treatment than pharmacotherapy (MD −20 minutes [95%CI, −39 to −2]). The combination was better than pharmacotherapy alone in the remission at post-treatment (OR 1.86 [95%CI, 1.11 to 3.09]) and wake after sleep onset at post-treatment (MD −11 minutes [95% CI, −22 to −1]). There was no clear evidence of difference between CBT-I and combination.

Sensitivity analyses generally confirmed the superiority of CBT-I over pharmacotherapy. The post hoc sensitivity analysis with a more stringent categorization suggested that treatment strategies starting with CBT-I (acute CBT-I followed by post-acute CBT-I, acute CBT-I followed post-acute pharmacotherapy, acute CBT-I followed by naturalistic follow-up) and starting with combination and then CBT-I alone were more beneficial than the treatment strategy starting with pharmacotherapy alone with naturalistic follow-up. (eAppendix7)

Given the weighted average proportion of remitters in pharmacotherapy-initiating arms in the long-term at 28%, we estimated that CBT-I led to remission in 41% (95%CI, 31% to 53%) and combination in 40% (95%CI, 25% to 56%) of the patients. The weighted average proportion of dropouts in pharmacotherapy-initiating arms in the long-term was estimated to be 39%. Using this number, we estimated that CBT-I led to dropouts in 21% (95%CI, 14% to 30%) and combination in 29% (95%CI, 17% to 45%) of the patients. (Table3)

## DISCUSSION

We performed the first systematic review and NMA of the initial treatment choices for insomnia, aiming to identify which treatment may maximize the chance of remission in the long-term. Our findings showed that starting with CBT-I was superior to starting with pharmacotherapy both in the long-term and at the end of the acute treatment phase, both in terms of efficacy and acceptability. Combination therapy may be more effective and acceptable than pharmacotherapy alone, but there was no evidence of its superiority over CBT-I alone. Total sleep time at post-treatment was shortest in CBT-I at post-treatment, but the difference was unclear in the long-term.

Based on these findings, we suggest people to start insomnia treatment with CBT-I alone. Combining sleep medication with CBT-I may be as effective as CBT-I alone, but it entails more cost and possible side effects, such as residual sedation[6] dependence/withdrawal,[30] and falls.[31] In the case of pregnant women, there is also an elevated risk for miscarriage.[32] However, given the short total sleep time in the CBT-I arms at post-treatment, patients who are vulnerable to sleep loss may prefer starting with combination or pharmacotherapy. Some may find CBT-I burdensome and prefer pharmacotherapy alone.

We confirmed the superiority of CBT-I and combination therapy over pharmacotherapy for patients with hypnotic-free insomnia at post-treatment as previously suggested in people with insomnia in general.[11] The previous network meta-analysis did not support the long-term superiority of any treatment over another because of the limited numbers of trials included.[11] This may be because they categorized the long-term follow-ups in three categories (1-3 months, 6-8 months, and 12-24 months) and lost the statistical power to detect a difference even though they all tended to favor CBT-I. We reasoned that comparative effectiveness was likely to remain stable in the long-term,[33] and therefore prespecified in the protocol to use the longest follow-up in 3-12 months for the long-term follow-up outcome. Another strength of our study is that we defined CBT-I as those including effective components[12] and pharmacotherapies as those shown effective[6] so that comparing them would be clinically relevant.

The clinical practice guideline of the American College of Physicians recommended CBT-I as the initial treatment for insomnia based on a series of pairwise meta-analyses of active versus control conditions that investigated the efficacy and safety profile of treatments.[7] Our findings further strengthen this recommendation by providing evidence on comparative efficacy and acceptability based on network meta-analyses. Given patients’ preference of non-pharmacological therapy over pharmacotherapy,[9] clinicians, policy makers and reimbursement bodies should take actions to make CBT-I more widely accessible, so that patients’ preferences can be respected in everyday practices.

Our study has several limitations. First, none of the trials used dual orexin receptor antagonists, and whether the findings apply to these new hypnotics has yet to be evaluated. However, given the relatively mild efficacy of dual orexin receptor antagonists in the short-term and its sparse long-term evidence,[6] CBT-I should remain the best initial treatment option until proven otherwise. Second, all the CBT-I programs were provided by therapists. Self-help CBT-I, such as internet CBT-I, may be one solution to scale up its availability, but it remains unclear whether its effectiveness is comparable to therapist-guide CBT-I. Third, although the study found that CBT-I and combination therapy are more beneficial than pharmacotherapy both in the long-term (median 24 weeks follow-up) and at post-treatment (median 8 weeks), the relative effectiveness in the shorter period (several days after initiating the treatment) remains unclear.

## CONCLUSION

We found evidence that initiating treatment for chronic insomnia in adults with CBT-I leads to more beneficial results compared with starting with pharmacotherapy alone. While combining CBT-I and pharmacotherapy might be more effective than pharmacotherapy alone, we did not find evidence that would justify the use of combination therapy over using CBT-I alone. Healthcare providers, policymakers, and insurers should make CBT-I more widely accessible, so that patients’ preferences can be respected in everyday practices.

### Patient and public involvement

There was no patient or public involvement in the development of this manuscript.

## Funding

Nothing.

## Conflict of interest

YF has received consultancy fee from Panasonic and lecture fee from Otsuka outside the submitted work. MS reports personal fee from SONY outside submitted work.

TAF reports personal fees from Boehringer-Ingelheim, Daiichi Sankyo, DT Axis, Kyoto University Original, Shionogi, SONY and UpToDate, and a grant from DT Axis and Shionogi, outside the submitted work; In addition, TAF has a patent 7448125, and a pending patent 2022-082495, and intellectual properties for Kokoro-app licensed to Mitsubishi-Tanabe.

MP wrote treatment manuals and books for CBT-I, teaches CBT-I, and is a founder of Hypknowledge LLC.

## Data Availability

Codes for all analyses will be available in a repository on GitHub (https://github.com/ykfrkw/W2I) after publication.

## Acknowledgements

The views expressed are those of the authors and not necessarily those of affiliated organizations.

## Registration

The protocol is prospectively registered in PROSPERO (CRD42024505519).

This research was prospectively registered (#2023318NIe), Ethical Committee, Faculty of Medicine, The University of Tokyo.

## Data access statement

YF had full access to all the data in the study and takes responsibility for the integrity of the data and the accuracy of the data analysis.

## Data sharing

Codes for all analyses are available in a repository on GitHub (https://github.com/ykfrkw/W2I).

## Notes

### Funding Statement

This study did not receive any funding

### Author Declarations

Databases such as PubMed, CENTRAL, PsycINFO and WHO ICTRP

