## Appendix for "Initial treatment choices for long term remission of insomnia disorder in adults: a systematic review and network meta-analysis"

### **1. PROTOCOL**

The protocol was prospectively registered in PROSPERO (CRD42023450720). Below is the protocol as of 20 January 2024.

First draft: 20 September 2023

Last edited: 20 January 2024

**Title: Initial treatment choices for insomnia disorder in adults: a protocol for a systematic review and network meta-analysis**

**REVIEW QUESTION**

What are the long-term comparative efficacy and acceptability of cognitive behavioral therapy for insomnia, pharmacotherapy, and their combination when used as the initial treatment for insomnia disorder in adults?

**BACKGROUND**

Insomnia is common and disabling.(Roth et al., 2011) Meta-analyses of randomized controlled trials have shown the effectiveness of cognitive behavioral therapy for insomnia(Furukawa et al., 2024) and some pharmacotherapies.(De_Crescenzo et al., 2022) A recent network meta-analysis (NMA) suggested potential superiority of CBT-I over pharmacotherapies and their combination over pharmacotherapies both at the end of the acute phase treatment and at follow-up(Zhang et al., 2022) However, the NMA included hypnotic-resistant insomniacs and this does not answer the clinical question of which treatment strategy to choose when starting to treat medication-naïve insomnia. In this study, we will explore the relative efficacy and acceptability of CBT-I, pharmacotherapy, and their combination as the initial treatment choice with the use of NMA, focusing on trials that randomized treatment-naïve insomniacs only.

**METHODS**

We will follow the Preferred Reporting Items for Systematic reviews and Meta-Analyses (PRISMA) guideline extension for NMA.(Hutton et al., 2015) We will register the study protocol on PROSPERO.

**Data sources**

**Criteria for considering studies for this review**

***Study design***

We will include all randomized controlled trials that compared CBT-I, pharmacotherapies, or their combination against each other in the treatment of hypnotic-free adults with chronic insomnia.

***Participants***

We will include studies of patients of both genders aged 18 years or older with insomnia either diagnosed according to formal diagnostic criteria (such as the Diagnostic and Statistical Manual of Mental Disorders, the International Classification of Diseases or the International Classification of Sleep Disorders) or judged so by clinical experts (e.g. presence of significant symptoms). The criteria need to include significant distress or daytime impairment. The effect of including studies without a formal diagnosis of insomnia will be tested in a sensitivity analysis. We will include patients with psychiatric or physical comorbidities. The effect of including such studies will be examined in a sensitivity analysis.

We will exclude trials if patients currently using prescription or over-the-counter sleep medications were included. We will include studies if patients discontinued the medications for a certain period before randomization.

***Interventions and controls***

We regard CBT-I as a psychotherapy involving any one of the following effective components: cognitive restructuring for insomnia, third wave components for insomnia(mindfulness and acceptance and commitment therapy), sleep restriction or stimulus control.(Furukawa et al., 2024) We will include drugs that were proven to be effective in the recent NMA. (benzodiazepines, doxylamine, eszopiclone, lemborexant, seltorexant, suvorexant, trazodone, zaleplon, zolpidem, zopiclone) (De_Crescenzo et al., 2022) Where multiple arms are reported in a single trial, we will include only the relevant arms.

**Search methods for identification of studies**

We will carry out a comprehensive literature search in PubMed, Cochrane Central Register of Controlled Trials and PsycINFO. We will use a combination of index and free terms of psychological treatments and insomnia with filters for randomized clinical trials. We will also search WHO International Clinical Trials Registry Platform. We will impose no date, language or publication status restriction. We will check the reference lists of identified studies and review articles for additional potentially eligible records.

**TABLE 3 Search strings for PubMed, Cochrane Central Register of Controlled Trials and PsycINFO**

| Database | Search strings |
| --- | --- |
| PubMed | ( "psychotherapy"[Mesh] OR "psychotherap*"[All Fields] OR "cognitive behavioural therapy"[All Fields] OR "cognitive behavioral therapy"[All Fields] OR "CBT"[All Fields] OR "CBTI"[All Fields] OR "CBT-I"[All Fields] OR "cognitive therapy"[All Fields] OR "behavioural therapy"[All Fields] OR "behavioral therapy"[All Fields] OR "cognitive restructuring"[All Fields] OR "third wave"[All Fields] OR "mindfulness"[All Fields] OR "acceptance and commitment"[All Fields] OR "sleep restriction"[All Fields] OR "stimulus control"[All Fields] )  AND  ("hypnotic"[All Fields] OR "sleep medication"[All Fields] OR "benzodiazepines"[Mesh] OR "benzodiazepine"[All Fields] OR "brotizolam"[All Fields] OR "diazepam"[All Fields] OR "estazolam"[All Fields] OR "flunitrazepam"[All Fields] OR "flurazepam"[All Fields] OR "haloxazolam"[All Fields] OR "loprazolam"[All Fields] OR "lorazepam"[All Fields] OR "lormetazepam"[All Fields] OR "nimetazepam"[All Fields] OR "nitrazepam"[All Fields] OR "quazepam"[All Fields] OR "rilmazafone"[All Fields] OR "temazepam"[All Fields] OR "triazolam"[All Fields] OR "trazodone"[All Fields] OR "eszopiclone"[All Fields] OR "zopiclone"[All Fields] OR "doxylamine"[All Fields] OR "zolpidem"[All Fields] OR "seltorexant"[All Fields] OR "lemborexant"[All Fields] OR "suvorexant"[All Fields] OR "zaleplon"[All Fields] )  AND  ("sleep initiation and maintenance disorders"[Mesh] OR "insomnia"[All Fields] )  AND  ("randomized controlled trial"[pt] OR "controlled clinical trial"[pt] OR randomized[tiab] OR placebo[tiab] OR "clinical trials as topic"[Mesh:NoExp] OR randomly[tiab] OR trial[ti] NOT (animals[Mesh] NOT humans[Mesh])) |
| Cochrane Central Register of Controlled Trials | ([mh psychotherapy] OR psychotherap* OR "cognitive behavioural therapy" OR "cognitive behavioral therapy" OR CBT OR CBTI OR CBT-I OR "cognitive therapy" OR "behavioural therapy" OR "behavioral therapy" OR "cognitive restructuring" OR "third wave" OR mindfulness OR "acceptance and commitment" OR "sleep restriction" OR "stimulus control")  AND  (hypnotic OR "sleep medication" OR [mh benzodiazepines] OR benzodiazepine OR brotizolam OR diazepam OR estazolam OR flunitrazepam OR flurazepam OR haloxazolam OR loprazolam OR lorazepam OR lormetazepam OR nimetazepam OR nitrazepam OR quazepam OR rilmazafone OR temazepam OR triazolam OR trazodone OR eszopiclone OR zopiclone OR doxylamine OR zolpidem OR seltorexant OR lemborexant OR suvorexant OR zaleplon)  AND  ([mh "sleep initiation and maintenance disorders"] OR insomnia) |
| PsycINFO via Ebsco | ((MH psychotherapy+) OR psychotherap* OR "cognitive behavioural therapy" OR "cognitive behavioral therapy" OR CBT OR CBTI OR CBT-I OR "cognitive therapy" OR "behavioural therapy" OR "behavioral therapy" OR "cognitive restructuring" OR "third wave" OR mindfulness OR "acceptance and commitment" OR "sleep restriction" OR "stimulus control")  AND  (hypnotic OR "sleep medication" OR (MH benzodiazepines+) OR benzodiazepine OR brotizolam OR diazepam OR estazolam OR flunitrazepam OR flurazepam OR haloxazolam OR loprazolam OR lorazepam OR lormetazepam OR nimetazepam OR nitrazepam OR quazepam OR rilmazafone OR temazepam OR triazolam OR trazodone OR eszopiclone OR zopiclone OR doxylamine OR zolpidem OR seltorexant OR lemborexant OR suvorexant OR zaleplon)  AND  ((MH "sleep initiation and maintenance disorders+") OR insomnia)  AND  (MH randomized controlled trials OR MH double-blind studies OR MH single-blind studies OR MH random assignment OR MH pretest-posttest design OR MH cluster sample OR TI (randomised OR randomized) OR AB (random*) OR TI (trial) OR (MH (sample size) AND AB (assigned OR allocated OR control)) OR MH (placebos) OR PT (randomized controlled trial) OR AB (control W5 group) OR MH (crossover design) OR MH (comparative studies) OR AB (cluster W3 RCT)) NOT ((MH animals+ OR MH animal studies OR TI (animal model*)) NOT MH human) |
| WHO International Clinical Trials Registry Platform | (psychotherap* OR "cognitive behavioural therapy" OR "cognitive behavioral therapy" OR "CBT" OR "CBTI" OR "CBT-I" OR "cognitive therapy" OR "behavioural therapy" OR "behavioral therapy" OR "cognitive restructuring" OR "third wave" OR mindfulness OR "acceptance and commitment" OR "sleep restriction" OR "stimulus control")  AND  (hypnotic OR "sleep medication" OR benzodiazepines OR benzodiazepine OR brotizolam OR diazepam OR estazolam OR flunitrazepam OR flurazepam OR haloxazolam OR loprazolam OR lorazepam OR lormetazepam OR nimetazepam OR nitrazepam OR quazepam OR rilmazafone OR temazepam OR triazolam OR trazodone OR eszopiclone OR zopiclone OR doxylamine OR zolpidem OR seltorexant OR lemborexant OR suvorexant OR zaleplon)  AND  ("sleep initiation and maintenance disorders " OR insomnia) |

**Data collection and analysis**

**Selection of studies**

Two review authors will independently screen titles and abstracts of all the potential studies we identify as a result of the search and code them as ‘retrieve’ or ‘do not retrieve’. We will retrieve the full text study reports/publications and two review authors will independently screen the full text and identify studies for inclusion and identify and record reasons for exclusion of the ineligible studies. We will resolve any disagreement through discussion or, if required, we will consult a third reviewer. We will identify publications from the same study so that each study rather than each report is the unit of analysis in the review. We will record the selection process in sufficient detail to complete a PRISMA flow diagram.

**Data items**

Two review authors will extract independently data from the included studies. Any disagreement will be resolved through discussion, or discussed with a third person if necessary. We will abstract the following information.

***1. Characteristics of the studies***

Name of the study, year of publication, country, study site (single or multi-center), recruitment, population characteristics (mean age, number of women, definition of insomnia), intervention, outcomes (scale used for the primary outcome)

***2. Risk of bias***

We will use Cochrane Risk of Bias 2.0 tool (RoB2) (Sterne et al., 2019) to assess the risk of bias of the primary outcome. We will report the inter-rater agreement in terms of percentage agreement and kappa.

***3. Data to calculate effect sizes***

We will extract data to calculate effect sizes (the number of patients randomized to each arm, the number of patients assessed, the number of remitters, the scale used, the mean, standard deviation and the number assessed for continuous outcomes) When only change from baseline to endpoint is reported for continuous outcomes, we will use it instead of endpoint mean.

**Primary outcome and secondary outcomes**

The primary outcome

1. Efficacy at long-term follow-up. (Remission defined as reaching a satisfactory state at endpoint measured by any validated self-reported scale. Dichotomous. Longest follow-up between 3 to 12 months)

Secondary outcomes are as follows;

2. Acceptability: dropouts for any reason at long-term follow-up (dichotomous)

3. Sleep diary measures at long-term follow-up (continuous)

3.1. Sleep efficiency at long-term follow-up (%)

3.2. Total sleep time at long-term follow-up (minutes)

3.3. Sleep latency at long-term follow-up (minutes)

3.4. Wake after sleep onset at long-term follow-up (minutes)

4. Efficacy at long-term follow-up (continuous)

5. Efficacy at post-treatment: remission defined as reaching a satisfactory state at endpoint measured by any validated self-reported scale (dichotomous)

6. Acceptability: dropouts for any reason at post-treatment (dichotomous)

7. Sleep diary measures at post-treatment (continuous)

7.1. Sleep efficiency at post-treatment (%)

7.2. Total sleep time at post-treatment (minutes)

7.3. Sleep latency at post-treatment (minutes)

7.4. Wake after sleep onset at post-treatment (minutes)

8. Efficacy at post-treatment (continuous)

Intention-to-treat analysis will be prioritized whenever available. We will use the number of participants randomized as the denominator for dichotomous outcomes. We will use odds ratio for dichotomous outcomes, mean difference for continuous outcomes expressed in minutes and percent.

**Hierarchy of outcome measures**

For efficacy, we will prioritize the remission using the Insomnia Severity Index (7 or less points at endpoint) and its imputed number. If it is not reported, we will use the following scales in this order: the remission using the Sleep Condition Indicator (17 or more points at endpoint); the remission using the Functional Outcomes of Sleep Questionnaire-10 (18 or more points at endpoint), and then its imputed number; remission using the Epworth Sleepiness Scale (10 or less points at endpoint), and then its imputed number; remission using the Pittsburgh Sleep Quality Index (5 or less points at endpoint), and then its imputed number; remission using the Athens Insomnia Scale (5 or less points at endpoint), and then its imputed number; remission using any other validated self-reported scales; remission using sleep diary measures (SE larger than 85% at endpoint; WASO less than 30 minutes at endpoint; SL less than 30 minutes at endpoint) and its imputed number.

When any of the measure is reported using another definition of remission than stated above, we will use the definition stated by the authors. When any of the measure is reported only in continuous values, we will impute remission using mean and standard deviation.

**Statistical analysis**

We will examine transitivity, an important underlying assumption of the network meta-analysis model, by creating a table of important trial and patient characteristics to see if potential effect modifiers (publication year, mean age, baseline severity) are similarly distributed among treatment comparisons. If we deem transitivity to be a plausible assumption, we will proceed with performing a network meta-analysis. Given the expected clinical and methodological heterogeneity of treatment effects among the studies, we will use the random-effects model, assuming a common heterogeneity parameter across the network.

Lack of transitivity may manifest as inconsistency in the network, in case there are closed loops. We will check for consistency using by comparing direct and indirect estimates and using global consistency test (design-by-treatment)(White et al., 2012). If the prerequisites of network meta-analysis are not met, or in case of large unexplained inconsistency, we will only present direct (i.e. from pairwise meta-analyses) and indirect evidence for each treatment comparison.

We will perform all analyses in R using *netmeta* package(Rücker et al., 2020) to conduct network meta-analysis and *meta* package(Balduzzi et al., 2019) to assess the reporting bias.

**Certainty of evidence**

We will assess the certainty of evidence in network estimates of the primary outcome using CINeMA.(Nikolakopoulou et al., 2020)

**Publication bias**

We will assess the presence of small study effects, including publication bias, in the evidence set with ten or more trials by examining asymmetry in the contour-enhanced funnel plots.

**Sensitivity analyses**

1. Excluding studies without formal diagnosis of insomnia

2. Excluding studies focusing on patients with comorbidities (both physical and psychological)

3. Excluding studies with overall high dropout rate (20% or more)

4. Excluding studies at high overall risk of bias

**Patient and public involvement**

There was no patient or public involvement in the development of this manuscript.

**Funding**

Self-funded

**Declaration of interests**

YF has received consultancy fee from Panasonic outside the submitted work.

MS reports personal fees from SONY outside the submitted work.

TAF reports personal fees from Boehringer-Ingelheim, DT Axis, Kyoto University Original, Shionogi and SONY, and a grant from Shionogi, outside the submitted work; In addition, TAF has patents 2020-548587 and 2022-082495 pending, and intellectual properties for Kokoro-app licensed to Mitsubishi-Tanabe.

MP wrote treatment manuals and books for CBT-I, teaches CBT-I, and is a founder of Hypknowledge LLC.

**Acknowledgements**

The views expressed are those of the authors and not necessarily those of affiliated organizations.

**Registration**

We will register the study protocol on PROSPERO.

**Amendments**

April 10, 2024. We decided to conduct two post-hoc sensitivity analyses to test the influence of arm definitions.

**Additional table. Interventions**

| Intervention | Description |
| --- | --- |
| Cognitive components | |
| Cognitive restructuring | Skills to identify, challenge and change unrealistic beliefs about sleep that may disturb sleep. Sometimes simply called cognitive therapy. |
| Third wave components | Mindfulness and acceptance and commitment therapy. Mindfulness is a form of meditation emphasizing a non-judgmental state of heightened or complete awareness of one’s thoughts, emotions, or experiences on a moment-to-moment basis. Acceptance and commitment therapy focuses on accepting the feelings and thoughts associated with insomnia through value-based behaviors. |
| Behavioral components | |
| Sleep restriction | Skills to improve sleep by limiting time in bed. First, time in bed is restricted to the average sleep duration and then it is increased or decreased depending on sleep efficiency. |
| Stimulus control | Skills to re-associate the bed with sleep. Patients are instructed to; wake up at the same time every morning, refrain from daytime napping, go to bed only when sleepy, get out of bed when unable to sleep, and use the bed/bedroom for sleep and sex only. |

### **2. SCREENIING PROCESS AND RESULTS**

#### FLOW DIAGRAM

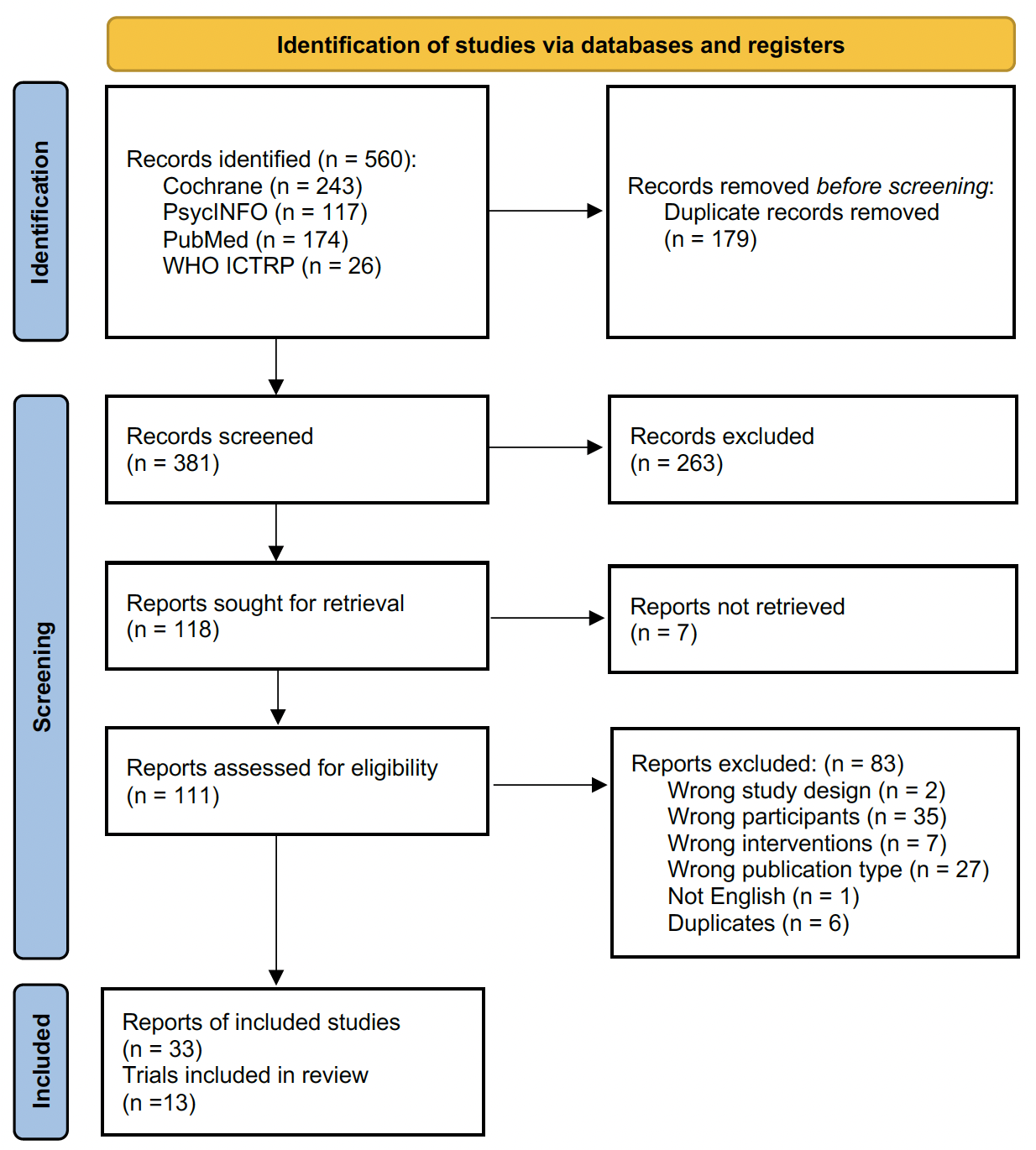

#### LIST OF EXCLUDED STUDIES WITH REASONS (EXAMPLES)

**Wrong participants (no mention of daytime impairment)**

Waters WF, Hurry MJ, Binks PG, et al. Behavioral and hypnotic treatments for insomnia subtypes. Behav Sleep Med. 2003;1(2):81-101. doi:10.1207/S15402010BSM0102_2

**Not hypnotic-naïve**

Ayabe N, Okajima I, Nakajima S, et al. Effectiveness of cognitive behavioral therapy for pharmacotherapy-resistant chronic insomnia: a multi-center randomized controlled trial in Japan. Sleep Med. 2018;50:105-112. doi:10.1016/j.sleep.2018.05.038

**Wrong drugs**

Li, X., Jiang, X., Liu, Y. et al. Efficacy of dexmedetomidine with cognitive behavioral therapy for treating chronic insomnia related to conditioned arousal: a randomized controlled trial. Sleep Biol. Rhythms 14, 75–85 (2016). https://doi.org/10.1007/s41105-015-0025-7

#### LIST OF ONGOING TRIALS

- ChiCTR2000034739
- CTRI/2020/12/029734
- jRCT2032230353
- NCT00044629
- NCT04468776

#### THE REVISED COCHRANE RISK OF BIAS FOR THE PRIMARY ANALYSIS

#### Risk of Bias 2 interpretation

We evaluated the risk of bias about the primary outcome, not the study quality, using the revised Cochrane risk-of-bias tool for randomized trials (RoB2). Here, we describe how we interpreted the signaling questions in each domain.

**Domain 1. Risk of bias arising from the randomization process**

1.1 Was the allocation sequence random?

1.2 Was the allocation sequence concealed until participants were enrolled and assigned to interventions?

We excluded studies where sequence generation was not clearly random, or where the allocation was clearly not concealed, were excluded.

1.3 Did baseline differences between intervention groups suggest a problem with the randomization process?

We evaluated if there were baseline differences in age, gender, and the primary outcome measure.

**Domain 2. Risk of bias due to deviations from intended interventions**

2.1. Were participants aware of their assigned intervention during the trial?

2.2. Were carers and people delivering the interventions aware of participants' assigned intervention during the trial?

In most cases, yes.

2.3. Were there deviations from the intended intervention that arose because of the trial context?

The term “trial context” refers to effects of recruitment and engagement activities on trial participants and when trial personnel undermine the implementation of the trial protocol in ways that would not happen outside the trial (e.g. daily practice). We expect non-adherence to the active treatment to occur outside the trials, too. Therefore we rated No.

2.6 Was an appropriate analysis used to estimate the effect of assignment to intervention?

We used ITT or mITT in the primary analysis so we rated Yes.

**Domain 3. Risk of bias due to missing outcome data**

3.1 Were data for this outcome available for all, or nearly all, participants randomized?

In the guidance document of RoB2, it is stated that the proportion required for dichotomous outcomes depends on the risk of the event. As we expect the control event rate of 20-30% and the experimental event rate of 40-60%, we decided to use 10% as the threshold.

3.2 Is there evidence that the result was not biased by missing outcome data?

We rated yes when appropriate sensitivity analyses were conducted and they showed the result was not likely to be biased by missing outcome data.

3.3 Could missingness in the outcome depend on its true value?

Yes.

3.4 Is it likely that missingness in the outcome depended on its true value?

We rated yes when the proportions of missing data differed significantly (larger than 10%) between the arms or when the reasons for missing differed between the arms.

**Domain 4. Risk of bias in measurement of the outcome**

4.1 Was the method of measuring the outcome inappropriate?

Probably not.

4.2 Could measurement or ascertainment of the outcome have differed between intervention groups?

Probably not.

4.3 Were outcome assessors aware of the intervention received by study participants?

Yes.

4.4 Could assessment of the outcome have been influenced by knowledge of intervention received?

We rated no when more than two active comparators were used.

**Domain 5. Risk of bias in selection of the reported result**

5.1 Were the data that produced this result analyzed in accordance with a pre-specified analysis plan that was finalized before unblinded outcome data were available for analysis?

Is the numerical result being assessed likely to have been selected, on the basis of the results, from...

5.2. ... multiple eligible outcome measurements within the outcome domain?

5.3 ... multiple eligible analyses of the data?

We predefined in the protocol the hierarchy of outcome measures. We rated low risk of bias for the domain 5 when the trial reported the top priority outcome (ISI remission), or it reported ISI (continuous value) in a way that predefined in the statistical analysis plan. In most cases, details of statistical analysis plans were unavailable and we therefore rated some concerns.

The overall risk of bias of the primary outcome for a study was judged “low” only when the study was judged to be at low risk of bias for ALL the five domains of potential biases arising from the randomization process, due to deviations from intended interventions, due to missing outcome data, in measurement of the outcome or selection of reported results, “some concerns” when one to three domains were judged to have some concerns and none at high risk, and “high” when the study was judged to be at high risk in one or more domains or to have some concerns in four or more domains

### **3. ASSESSMENT OF TRANSITIVITY**

#### Box plots

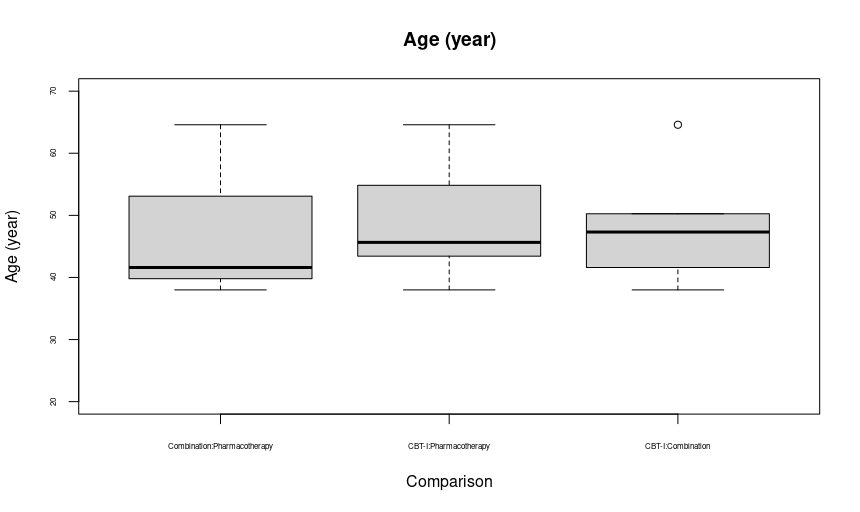

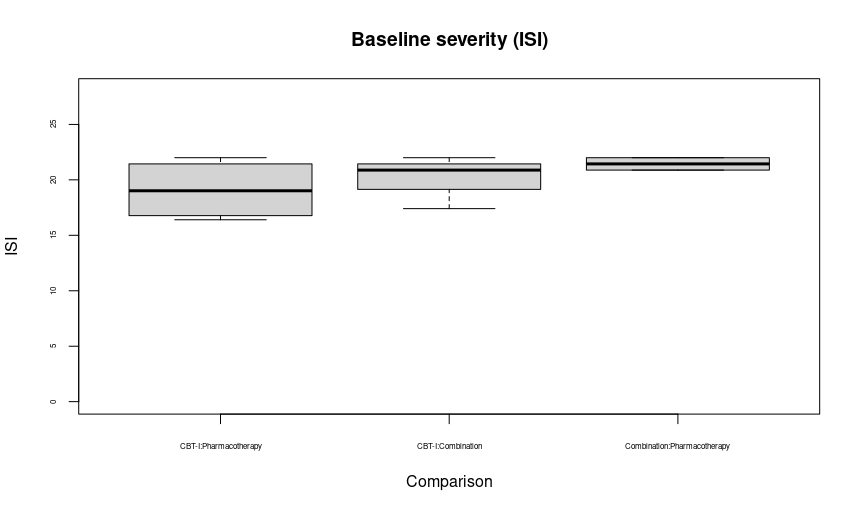

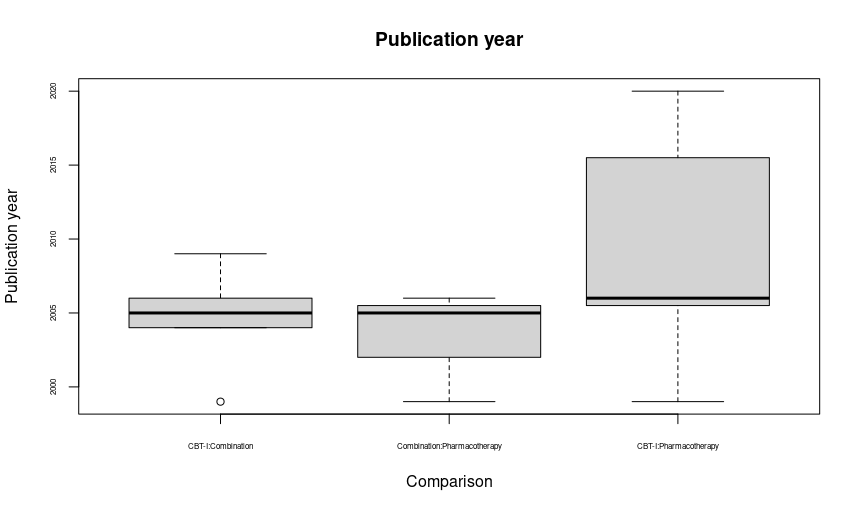

#### Global (design-by-treatment) test

```
> decomp.design(net_nma)
Q statistics to assess homogeneity / consistency

                    Q      df   p-value
Total              10.44  10  0.4032
Within designs    4.68   8  0.7916
Between designs   5.76   2  0.0561

Design-specific decomposition of within-designs Q statistic

                            Design   	 Q    df  p-value
 Pharmacotherapy:CBT-I:Combination 	2.80  4  0.5920
             Pharmacotherapy:CBT-I	1.87  3  0.6000
                 CBT-I:Combination 	0.01  1  0.9306

Between-designs Q statistic after detaching of single designs
(influential designs have p-value markedly different from 0.0561)

                   Detached design    Q df p-value
             Pharmacotherapy:CBT-I 3.56  1  0.0591
                 CBT-I:Combination 3.80  1  0.0513
 Pharmacotherapy:CBT-I:Combination 0.00  0      --

Q statistic to assess consistency under the assumption of
a full design-by-treatment interaction random effects model

                   Q     df  p-value tau.within tau2.within
Between designs 5.76  2   0.0561          0           0
```

#### Local (back-calculation) test

```
> netsplit(net_nma)
Separate indirect from direct evidence (SIDE) using back-calculation method

Random effects model:

comparison 		k prop    nma direct indir.     RoR    z	  p-value
CBT-I:Combination	 	5 0.96 1.0658 1.1577 0.1385  8.3595 1.54  0.1230
CBT-I:Pharmacotherapy 	7 0.98 1.8176 1.9032 0.1642 11.5874 1.42  0.1550
Combination:Pharmacotherapy 	3 0.32 1.7054 2.2528 1.4940  1.5079 0.57  0.5688

Legend:
 comparison - Treatment comparison
 k          - Number of studies providing direct evidence
 prop       - Direct evidence proportion
 nma        - Estimated treatment effect (OR) in network meta-analysis
 direct     - Estimated treatment effect (OR) derived from direct evidence
 indir.     - Estimated treatment effect (OR) derived from indirect evidence
 RoR        - Ratio of Ratios (direct versus indirect)
 z          - z-value of test for disagreement (direct versus indirect)
 p-value    - p-value of test for disagreement (direct versus indirect)
```

#### Comparing τ2 against empirical distributions

```
> d1=rnorm(1000000, mean = -1.09, sd= 1.27)
> quantile(exp(d1), c(0.0206,0.5, .975))
     2.06%        50%      97.5%
0.02525211 0.33717870 4.06069176
> mean(d1>log(0.0206))
[1] 0.986145
```

#### Prediction intervals

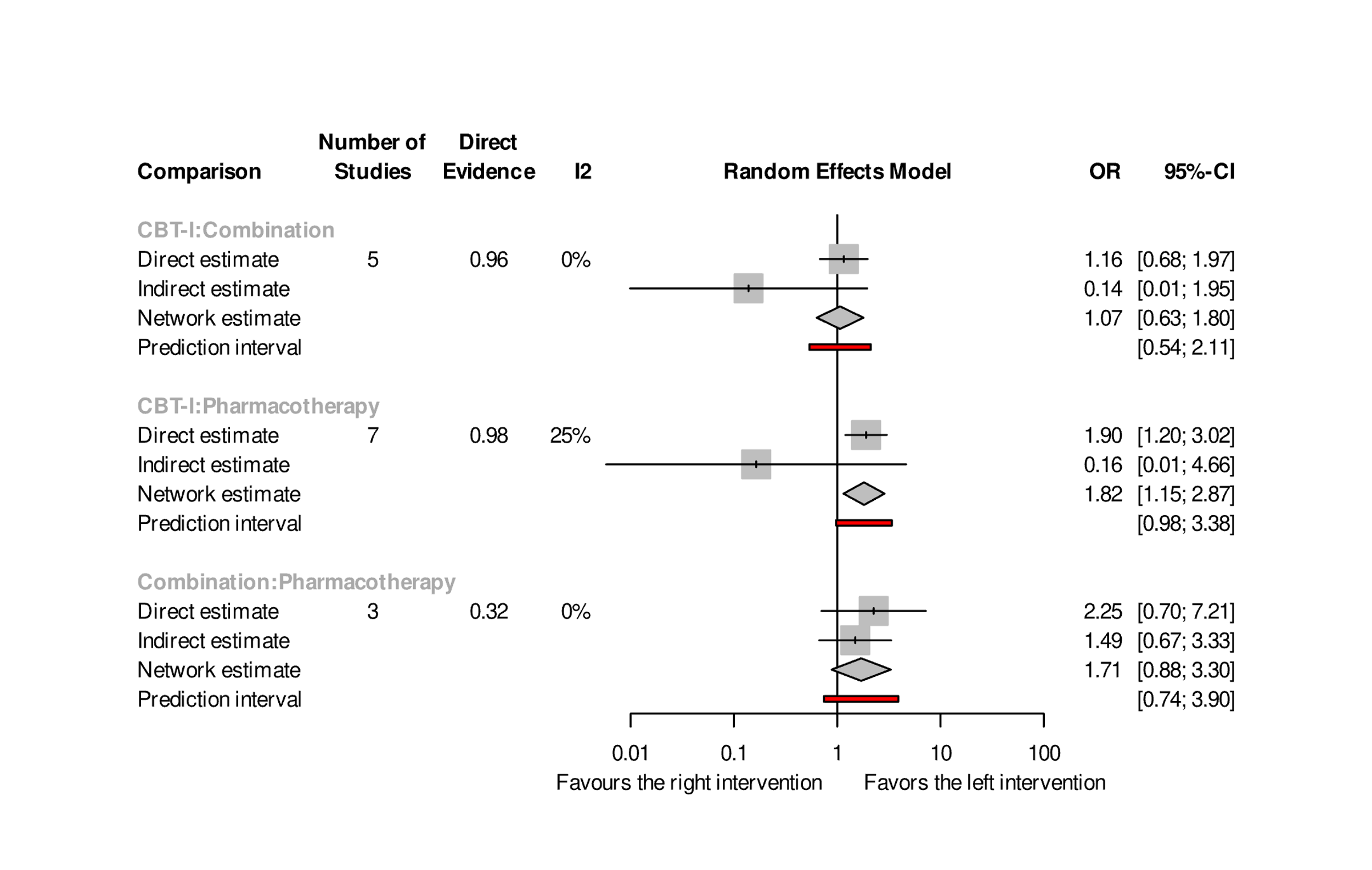

### **4. LEAGUE TABLES**

CBT-I: cognitive-behavioral therapy for insomnia. Figures represents the odds ratios. The bottom-left part shows the results of network meta-analysis and the upper-right part the results of pairwise meta-analyses.

Remission in the long-term (primary)

| CBT-I | 1.16 (0.68 to 1.97) | 1.90 (1.20 to 3.02) |
| --- | --- | --- |
| 1.07 (0.63 to 1.80) | Combination | 2.25 (0.70 to 7.21) |
| 1.82 (1.15 to 2.87) | 1.71 (0.88 to 3.30) | Pharmacotherapy |

Dropout in the long-term

| CBT-I | 0.55 (0.29 to 1.03) | 0.44 (0.27 to 0.70) |
| --- | --- | --- |
| 0.64 (0.36 to 1.16) | Combination | 0.38 (0.12 to 1.23) |
| 0.42 (0.26 to 0.66) | 0.64 (0.32 to 1.28) | Pharmacotherapy |

Remission at post-treatment

| CBT-I | 0.91 (0.56 to 1.46) | 1.56 (1.04 to 2.35) |
| --- | --- | --- |
| 0.86 (0.55 to 1.33) | Combination | 2.10 (1.06 to 4.17) |
| 1.59 (1.07 to 2.35) | 1.86 (1.11 to 3.09) | Pharmacotherapy |

Dropout at post-treatment

| CBT-I | 0.53 (0.20 to 1.41) | 0.50 (0.28 to 0.89) |
| --- | --- | --- |
| 0.72 (0.30 to 1.72) | Combination | 0.39 (0.09 to 1.65) |
| 0.47 (0.27 to 0.83) | 0.66 (0.26 to 1.69) | Pharmacotherapy |

### **5. ASSESSMENT OF PUBLICATION BIAS AND SMALL STUDY EFFECTS**

We did not evaluate publication bias and small-study effects using funnel plots due to the limited number of trials.

### **6. Confidence in Network Meta-Analysis (CINeMA)**

We considered reporting bias of all the comparisons to be “low.” We regarded odds ratio of 1.5 as clinically important effect size. We downgraded the comparisons with number of studies less than 3. We downgraded one level with one domain of “major concerns” and two or three domains of “some concerns.”

| **Comparison** | **k** | **Within-study bias** | **Reporting bias** | **Indirectness** | **Imprecision** | **Heterogeneity** | **Incoherence** | **Confidence** | **Reason(s) for  downgrading** |
| --- | --- | --- | --- | --- | --- | --- | --- | --- | --- |
| **CBT-I:Combination** | 5 | Some concerns | Low risk | No concerns | Major concerns | No concerns | No concerns | Moderate | ["Imprecision"] |
| **CBT-I:Pharmacotherapy** | 7 | No concerns | Low risk | No concerns | No concerns | Some concerns | No concerns | High |  |
| **Combination:Pharmacotherapy** | 3 | Some concerns | Low risk | No concerns | Some concerns | No concerns | No concerns | Moderate | ["Within-study bias", "Imprecision"] |

### **7. SENSITIVITY ANALYSES**

#### S1. Excluding studies without formal diagnosis of insomnia　(Excluding Vgotzas2020)

```
Number of studies: k = 8
Number of pairwise comparisons: m = 14
Number of treatments: n = 3
Number of designs: d = 3

Random effects model

Treatment estimate (sm = 'OR', comparison: other treatments vs 'Pharmacotherapy'):
                    OR           	95%-CI   		 z 	p-value
CBT-I           1.9406 	[1.1223; 3.3556] 	2.37  	0.0176
Combination    1.7414 	[0.8232; 3.6836] 	1.45  	0.1468
Pharmacotherapy      .                .    .       .

Quantifying heterogeneity / inconsistency:
tau^2 = 0.0739; tau = 0.2718; I^2 = 13.6% [0.0%; 54.9%]

Tests of heterogeneity (within designs) and inconsistency (between designs):
                  	  Q 	d.f. 	p-value
Total          	 10.42    	9  	0.3179
Within designs   	4.57    	7  	0.7122
Between designs 	 5.85    	2  	0.0538
```

#### S2. Excluding studies focusing on patients with comorbidities

We could not perform this pre-specified sensitivity analysis due to the lack of trials.

#### S3. Excluding studies with overall high dropout rate (20% or more)　(Excluding Jacobs2004, Morin1999, Morin2009, Morin2020, Siversten2006, Vallieres2005, Vgotzas2020)

```
Number of studies: k = 2
Number of pairwise comparisons: m = 4
Number of treatments: n = 3
Number of designs: d = 2

Random effects model

Treatment estimate (sm = 'OR', comparison: other treatments vs 'Pharmacotherapy'):
                    OR   	95%-CI    		z 	p-value
CBT-I           3.0317 	[0.4778; 19.2366] 	1.18  	0.2394
Combination   2.3196 	[0.2025; 26.5646] 	0.68  	0.4988
Pharmacotherapy      .                 .    .       .

Quantifying heterogeneity / inconsistency:
tau^2 = 1.0878; tau = 1.0430; I^2 = 61.1% [0.0%; 91.0%]

Tests of heterogeneity (within designs) and inconsistency (between designs):
                   	Q 	d.f. 	p-value
Total         	2.57    	1  	0.1089
Within designs  	0.00    	0      	--
Between designs 	2.57    	1  	0.1089
```

#### S4. Excluding studies at high overall risk of bias (Excluding Jacobs2004, Siversten2006, Wu2006)

```
Number of studies: k = 6
Number of pairwise comparisons: m = 10
Number of treatments: n = 3
Number of designs: d = 3

Random effects model

Treatment estimate (sm = 'OR', comparison: other treatments vs 'Pharmacotherapy'):
                    OR  	95%-CI    	     z 	p-value
CBT-I            1.4714   [0.9301; 2.3277]   1.65 	0.0989
Combination     1.4662   [0.7241; 2.9690]   1.06  	0.2877
Pharmacotherapy      .                .  	  .        	.

Quantifying heterogeneity / inconsistency:
tau^2 = 0; tau = 0; I^2 = 0% [0.0%; 70.8%]

Tests of heterogeneity (within designs) and inconsistency (between designs):
                	   Q    d.f. p-value
Total             	 5.58    6  0.4723
Within designs    	 1.99    4  0.7382
Between designs  	 3.59    2  0.1662
```

#### S5. Excluding arms with switching (excluding CBT-I -> com, CBT-I -> pha, pha -> CBT-I, pha -> com) (Post hoc)

```
Number of studies: k = 9
Number of pairwise comparisons: m = 13
Number of treatments: n = 3
Number of designs: d = 3

Random effects model

Treatment estimate (sm = 'OR', comparison: other treatments vs 'Pharmacotherapy'):
                    OR           95%-CI    z p-value
CBT-I           2.0599 [1.1832; 3.5865] 2.55  0.0106
Combination     1.9255 [0.9224; 4.0195] 1.74  0.0810
Pharmacotherapy      .                .    .       .

Quantifying heterogeneity / inconsistency:
tau^2 = 0.0341; tau = 0.1846; I^2 = 6.6% [0.0%; 64.9%]

Tests of heterogeneity (within designs) and inconsistency (between designs):
                   Q     d.f. p-value
Total               9.64    9  0.3807
Within designs       2.72    7  0.9100
Between designs      6.92   2  0.0314
```

#### S6. Categorizing arms using both the initial and the second-step treatments (Post hoc)

```
Number of studies: k = 9
Number of pairwise comparisons: m = 25
Number of treatments: n = 10
Number of designs: d = 7

Random effects model

Treatment estimate (sm = 'OR', comparison: other treatments vs 'Pharmacotherapy -> nat'):
                                       OR            95%-CI    z p-value
CBT-I -> CBT-I                     3.5151 [1.1198; 11.0341] 2.15  0.0313
CBT-I -> nat                       3.6785 [1.6398;  8.2519] 3.16  0.0016
CBT-I -> Pharmacotherapy           4.2845 [1.1301; 16.2440] 2.14  0.0324
Combination -> CBT-I               6.0669 [1.8633; 19.7535] 2.99  0.0028
Combination -> Combination         2.7663 [0.8168;  9.3690] 1.63  0.1021
Combination -> nat                 2.2273 [0.8031;  6.1774] 1.54  0.1239
Pharmacotherapy -> CBT-I           2.8838 [0.7684; 10.8221] 1.57  0.1165
Pharmacotherapy -> Combination     5.5938 [0.4971; 62.9494] 1.39  0.1633
Pharmacotherapy -> nat                  .                 .    .       .
Pharmacotherapy -> Pharmacotherapy 2.6744 [0.7632;  9.3715] 1.54  0.1241

Quantifying heterogeneity / inconsistency:
tau^2 = 0; tau = 0; I^2 = 0% [0.0%; 67.6%]

Tests of heterogeneity (within designs) and inconsistency (between designs):
                   Q d.f. p-value
Total           5.43    7  0.6081
Within designs  1.45    3  0.6932
Between designs 3.97    4  0.4096
```

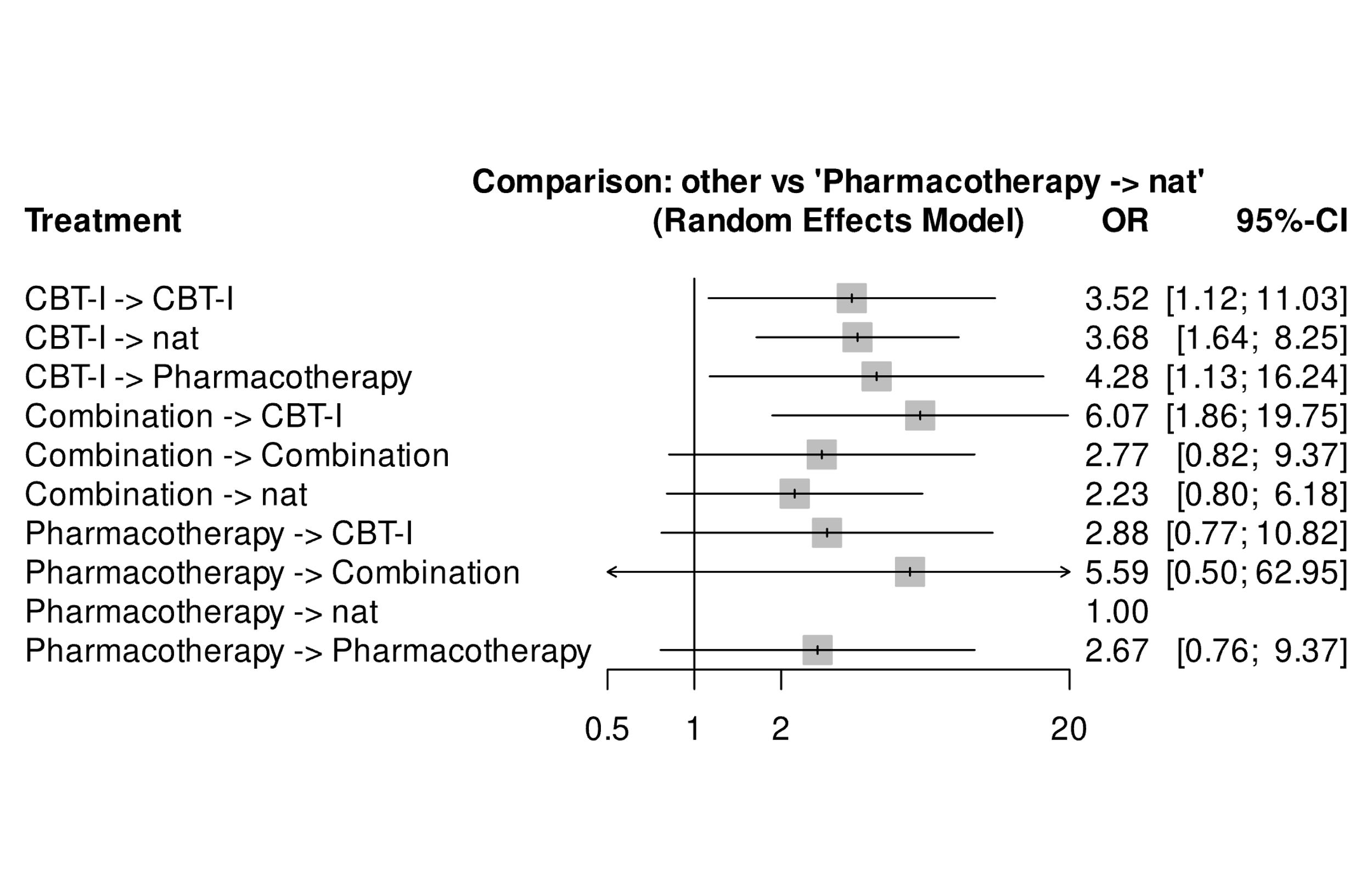

CBT-I=cognitive-behavioral therapy for insomnia; CI=confidence interval; nat=naturalistic follow-up; OR=odds ratio.

### **8. PRISMA-NMA**

| **Section/Topic** | **Item #** | **Checklist Item** | **Reported on Page #** |
| --- | --- | --- | --- |
| **TITLE** |  |  |  |
| Title | 1 | Identify the report as a systematic review *incorporating a network meta-analysis (or related form of meta-analysis).* | Title page |
| **ABSTRACT** |  |  |  |
| Structured summary | 2 | Provide a structured summary including, as applicable:  **Background:** main objectives  **Methods:** data sources; study eligibility criteria, participants, and interventions; study appraisal; and *synthesis methods, such as network meta-analysis.*  **Results:** number of studies and participants identified; summary estimates with corresponding confidence/credible intervals; *treatment rankings may also be discussed. Authors may choose to summarize pairwise comparisons against a chosen treatment included in their analyses for brevity.*  **Discussion/Conclusions:** limitations; conclusions and implications of findings.  **Other:** primary source of funding; systematic review registration number with registry name. | Abstract |
| **INTRODUCTION** |  |  |  |
| Rationale | 3 | Describe the rationale for the review in the context of what is already known*, including mention of why a network meta-analysis has been conducted.* | Introduction |
| Objectives | 4 | Provide an explicit statement of questions being addressed, with reference to participants, interventions, comparisons, outcomes, and study design (PICOS). | Introduction |
| **METHODS** |  |  |  |
| Protocol and registration | 5 | Indicate whether a review protocol exists and if and where it can be accessed (e.g., Web address); and, if available, provide registration information, including registration number. | Methods, eAppendix |
| Eligibility criteria | 6 | Specify study characteristics (e.g., PICOS, length of follow-up) and report characteristics (e.g., years considered, language, publication status) used as criteria for eligibility, giving rationale. *Clearly describe eligible treatments included in the treatment network, and note whether any have been clustered or merged into the same node (with justification).* | Methods |
| Information sources | 7 | Describe all information sources (e.g., databases with dates of coverage, contact with study authors to identify additional studies) in the search and date last searched. | Methods |
| Search | 8 | Present full electronic search strategy for at least one database, including any limits used, such that it could be repeated. | eAppendix |
| Study selection | 9 | State the process for selecting studies (i.e., screening, eligibility, included in systematic review, and, if applicable, included in the meta-analysis). | eAppendix |
| Data collection process | 10 | Describe method of data extraction from reports (e.g., piloted forms, independently, in duplicate) and any processes for obtaining and confirming data from investigators. | Methods |
| Data items | 11 | List and define all variables for which data were sought (e.g., PICOS, funding sources) and any assumptions and simplifications made. | eAppendix |
| **Geometry of the network** | **S1** | Describe methods used to explore the geometry of the treatment network under study and potential biases related to it. This should include how the evidence base has been graphically summarized for presentation, and what characteristics were compiled and used to describe the evidence base to readers. | Figure2 |
| Risk of bias within individual studies | 12 | Describe methods used for assessing risk of bias of individual studies (including specification of whether this was done at the study or outcome level), and how this information is to be used in any data synthesis. | eAppendix |
| Summary measures | 13 | State the principal summary measures (e.g., risk ratio, difference in means). *Also describe the use of additional summary measures assessed, such as treatment rankings and surface under the cumulative ranking curve (SUCRA) values, as well as modified approaches used to present summary findings from meta-analyses.* | Methods |
| Planned methods of analysis | 14 | Describe the methods of handling data and combining results of studies for each network meta-analysis. This should include, but not be limited to:   - *Handling of multi-arm trials;* - *Selection of variance structure;* - *Selection of prior distributions in Bayesian analyses; and* - *Assessment of model fit.* | Methods |
| **Assessment of Inconsistency** | **S2** | Describe the statistical methods used to evaluate the agreement of direct and indirect evidence in the treatment network(s) studied. Describe efforts taken to address its presence when found. | Results, eAppendix |
| Risk of bias across studies | 15 | Specify any assessment of risk of bias that may affect the cumulative evidence (e.g., publication bias, selective reporting within studies). | eAppendix |
| Additional analyses | 16 | Describe methods of additional analyses if done, indicating which were pre-specified. This may include, but not be limited to, the following:   - Sensitivity or subgroup analyses; - Meta-regression analyses; - *Alternative formulations of the treatment network; and* - *Use of alternative prior distributions for Bayesian analyses (if applicable).* | eAppendix |
| **RESULTS†** |  |  |  |
| Study selection | 17 | Give numbers of studies screened, assessed for eligibility, and included in the review, with reasons for exclusions at each stage, ideally with a flow diagram. | Appendix |
| **Presentation of network structure** | **S3** | Provide a network graph of the included studies to enable visualization of the geometry of the treatment network. | Figure1 |
| **Summary of network geometry** | **S4** | Provide a brief overview of characteristics of the treatment network. This may include commentary on the abundance of trials and randomized patients for the different interventions and pairwise comparisons in the network, gaps of evidence in the treatment network, and potential biases reflected by the network structure. | Figure2 |
| Study characteristics | 18 | For each study, present characteristics for which data were extracted (e.g., study size, PICOS, follow-up period) and provide the citations. | eAppendix |
| Risk of bias within studies | 19 | Present data on risk of bias of each study and, if available, any outcome level assessment. | eAppendix |
| Results of individual studies | 20 | For all outcomes considered (benefits or harms), present, for each study: 1) simple summary data for each intervention group, and 2) effect estimates and confidence intervals. *Modified approaches may be needed to deal with information from larger networks.* | NA |
| Synthesis of results | 21 | Present results of each meta-analysis done, including confidence/credible intervals. *In larger networks, authors may focus on comparisons versus a particular comparator (e.g. placebo or standard care), with full findings presented in an appendix. League tables and forest plots may be considered to summarize pairwise comparisons.* If additional summary measures were explored (such as treatment rankings), these should also be presented. | Figure2, Figure3 |
| **Exploration for inconsistency** | **S5** | Describe results from investigations of inconsistency. This may include such information as measures of model fit to compare consistency and inconsistency models, *P* values from statistical tests, or summary of inconsistency estimates from different parts of the treatment network. | eAppendix |
| Risk of bias across studies | 22 | Present results of any assessment of risk of bias across studies for the evidence base being studied. | eAppendix CINeMA |
| Results of additional analyses | 23 | Give results of additional analyses, if done (e.g., sensitivity or subgroup analyses, meta-regression analyses*, alternative network geometries studied, alternative choice of prior distributions for Bayesian analyses,* and so forth). | eAppendix |
| **DISCUSSION** |  |  |  |
| Summary of evidence | 24 | Summarize the main findings, including the strength of evidence for each main outcome; consider their relevance to key groups (e.g., healthcare providers, users, and policy-makers). | Discussion |
| Limitations | 25 | Discuss limitations at study and outcome level (e.g., risk of bias), and at review level (e.g., incomplete retrieval of identified research, reporting bias). *Comment on the validity of the assumptions, such as transitivity and consistency. Comment on any concerns regarding network geometry (e.g., avoidance of certain comparisons).* | Discussion |
| Conclusions | 26 | Provide a general interpretation of the results in the context of other evidence, and implications for future research. | Discussion |
| **FUNDING** |  |  |  |
| Funding | 27 | Describe sources of funding for the systematic review and other support (e.g., supply of data); role of funders for the systematic review. This should also include information regarding whether funding has been received from manufacturers of treatments in the network and/or whether some of the authors are content experts with professional conflicts of interest that could affect use of treatments in the network. | Funding |
